# Reactivity to neural tissue epitopes, aquaporin 4 and heat shock protein 60 is associated with activated immune-inflammatory pathways and the onset of delirium following hip fracture surgery

**DOI:** 10.1101/2022.06.24.22276844

**Authors:** Michael Maes, Paul Thisayakorn, Yanin Thipakorn, Saran Tantavisut, Sunee Sirivichayakul, Aristo Vojdani

## Abstract

**Objectives:** Activation of the immune-inflammatory response system (IRS) and a deficiency in the compensatory immunoregulatory system (CIRS), neuronal injuries, and alterations in the glutamate receptor (GlutaR), aquaporin-4 (AQP4), and heat shock protein 60 (HSP60) are involved in delirium. Increased serum levels of neurofilament protein (NFP), glial fibrillary acidic protein (GFAP) and myelin basic protein (MBP) are biomarker of neuronal injury and post-surgery cognitive impairments. Polyreactive antibodies (PAbs) contribute to the development of immune-mediated disease. This investigation delineates whether elevated IgA and IgG reactivity against those self-antigens is associated with enhanced IRS responses and delirium severity.

**Methods:** We measured peak Delirium Rating Scale (DRS) scores on days 2 and 3 following surgery in 59 hip fracture older adults, and IgA/IgG antibody levels against MBP, NFP, GFAP and myelin oligodendrocyte glycoprotein (MOG), GlutaR, N-Methyl-D-Aspartate receptor (NMDAR), APQ4 and HSP60.

**Results:** The IgA antibody levels against those self-antigens, especially GFAP, MBP and HSP60, strongly predict peak DRS scores on days 2 and 3 post-surgery. IgA reactivity against NMDAR and baseline DRS scores explained 40.6% of the variance in peak DRS scores, whilst IgA against NMDAR, IgG against MBP and age explained 29.1% of the variance in the IRS/CIRS ratio. There was no correlation between DRS scores and IgG directed against these self-antigens.

**Conclusions:** Increased IgA levels against neuronal self-antigens, AQP4, and HSP60 are risk factors for delirium. PAb-associated breakdown of immune tolerance, IRS activation and injuries in the neuronal cytoskeleton, oligodendrocytes, astrocytes, glial cells, and myelin sheath are involved in the pathophysiology of delirium.

## Background

Delirium is a neuropsychiatric syndrome characterized by impairments in neurocognitive functions, awareness, psychomotor functions and circadian rhythms, and is common in hospitalized older adults following hip fracture surgery [1, 2]. The prevalence of this condition in the post-operative period is 16.9-24% [3, 4]. A significant proportion of older adults who develop delirium are at increased risk of prolonged intensive care unit stay, increased medical comorbidities, and increased morbidity and mortality [5–7]. Risk factors of delirium comprise age, the aging brain, dysfunctions in neurotransmitters (serotonin, dopamine, glutamate, acetylcholine) and neuronal circuits, disorders in circadian rhythms, endocrine disorders, previous medical disorders including depression, stroke, and dementia [1,2,8,9,10] and duration of surgery and blood loss during surgery [2,11,12].

There is now evidence that delirium in older adults after surgery for hip fracture is associated with activation of immune-inflammatory pathways. Previous research has linked increases in peripheral C-reactive protein (CRP), interleukin-6 (IL-6), CXCL8 (IL-8), and tumor necrosis factor (TNF)-α to the onset of post-operative delirium [2]. These results indicate that activation of M1 macrophages and an acute phase (or inflammatory) response are associated with delirium. Another study found higher levels of IL-2, a T helper-1 (Th-1) cytokine, in delirium, along with higher levels of IL-1 and IL-6, but not higher levels of TNF-α, CXCL8, or CRP [2, 13]. Moreover, different studies showed an increased neutrophil/lymphocyte ratio (NLR) in patients with delirium [12,14–16], indicating activation of immune-inflammatory pathways [2].

Recently, we established that post-surgery delirium due to hip fracture is characterized by an increased activity of the immune-inflammatory response system (IRS) and a relative decrease in the activity of the compensatory immunoregulatory system (CIRS) as indicated by an increased IRS/CIRS ratio [2]. The IRS response comprises activation of the M1 phenotype (with increased levels of IL-6, CXCL8, and TNF-α), Th-1 phenotype (based on assays of IL-2, IL-12 ad interferon-γ (IFN-γ)), as well as the Th-17 phenotype (including IL-6 and IL-17) [2, 17]. The CIRS is the aggregate of different negative immunoregulatory factors, including Th-2 (e.g. IL-4), T regulatory (Treg) (e.g. IL-10) and cytokine receptors, e.g., the IL-1 receptor antagonist (sIL-1RA) which have anti-inflammatory properties thereby preventing hyperinflammation [2, 17]. In delirium, the IRS/CIRS ratio is increased indicating that CIRS activities (particularly IL-4 and sIL-1RA) are insufficient to regulate and attenuate the IRS [2].

Such findings suggest that delirium following hip fracture surgery is caused by an immune-inflammatory process initiated by hip tissue trauma and the release of damage-associated molecular patterns (DAMPs), which initiate a cascade of early inflammatory mechanisms ranging from local tissue injury to inflammatory cell activation to a peripheral IRS and neuroinflammation, and that these pathways are more pronounced in subjects with lower CIRS functions [2]. Nevertheless, there are no reports on the type of DAMPS that could be involved as for example heat shock proteins (HSP) which may act as DAMPS or perhaps “DAMPERs” [18, 19].

Furthermore, there is some evidence that dysfunctions in the neuroaxis and neuronal injuries are risk factors for acute brain failure [1, 10]. Hepatic failure, alcohol withdrawal, and ischemia, for example, can all cause glutamate excitotoxicity, resulting in acute brain failure. Anti-NMDA receptor (NMDAR) encephalitis (AMDARE) may cause psychomotor agitation, disinhibition, manic delirium, and catatonia [20], while lowering glutamate excitotoxicity reduces catatonia [21]. Another risk factor for delirium is neurofilament light, an indicator of neuronal injury, which is linked to neurodegenerative processes in the brain and delirium severity independently of inflammatory biomarkers [22, 23]. In addition, plasma glial fibrillary acid protein (GFAP), which is expressed in astrocytes and is involved in cell communication, is another putative biomarker of post-surgery cognitive impairments and neuronal injury [24]. In another investigation, immunological responses to brain damage markers such as myelin basic protein (MBP) were studied in alcohol withdrawal delirium [25]. Three days following the onset of delirium, antibodies to MBP were higher in patients than in controls, according to the latter investigation. Human aquaporin 4 (AQP4) water channels are detected in endothelial cells, the blood-brain barrier (BBB) and astrocytes and upregulation of AQP4 is related to dehydration and cerebral edema, both of which are risk factors for delirium [26, 27].

Following BBB and neuronal injuries, proteins such as AQP4, MBP and myelin oligodendrocyte glycoprotein (MOG), a transmembrane protein that regulates myelination and is expressed only by oligodendrocytes, may translocate to the bloodstream and mount immunoglobin (Ig) IgA/IgG antibody production against these antigens [25]. Moreover, polyreactive antibodies (PABs), including IgA PABs, may have homeostatic and immunoregulatory effects, but can be accompanied by breakdown of immune tolerance and PAB-related disorders [28]. In spite of this, there is no evidence that IgA/IgG reactivity against HSP60, glutamate receptor (GlutaR), NMDAR, MBP, NFP, GFAP, MOG, or AQP4 are linked with the onset or severity of delirium or the elevated IRS/CIRS ratio in delirium.

Hence, the present study was conducted to examine whether IgA/IgG to HSP60, GluR, NMDAR, MBP, MOG, NFP, AFAP and AQP4 are associated with the onset and severity of delirium and enhancement of IRS (increased IRS/CIRS ratio) in older adults with hip fracture surgery. According to our a priori hypothesis, delirium severity and the IRS response during delirium are characterized by increased IgA/IgG to HSP60 and neuronal self-antigens, implying that these indicators of immune and neuronal injuries are other risk factors for delirium.

## Methods

### Participants

Between June 2019 and February 2020, we recruited 59 older adults with hip fractures who were admitted to the Hip Fracture Pathway Inpatient Care at King Chulalongkorn Memorial Hospital in Bangkok, Thailand. The study included patients aged 65 and older who had a low energy impact hip fracture, underwent a hip fracture operation and were postoperatively transferred to the surgery intensive care unit (SICU) or orthopedic units. Exclusion criteria included: coma, major psychiatric illness (such as psycho-organic disorders, schizophrenia, bipolar disorders), a life-time history of (neuro)-inflammatory and/or neurodegenerative disease (including Parkinson’s disease, Alzheimer’s disease, multiple sclerosis), (auto)immune disorders (including inflammatory bowel disease, psoriasis, rheumatoid arthritis, systemic lupus erythematosus), a high energy impact hip fracture, intracranial vascular lesions, premorbid dementia, metastatic fractures, traumatic brain injury from falling. Patients with major depression who have been remitted, subjects with mild cognitive impairment, stroke patients one year after their acute stroke, could be included.

### Clinical assessments

Initially, demographic and clinical data from the research participants’ electronic medical records and bedside interviews were extracted. Within 24 hours of the surgery date, the baseline cognitive status and delirium scores were assessed. The cognitive status, delirium severity, and diagnosis were then re-assessed daily for three consecutive days postoperatively. The Delirium Rating Scale, Revised-98-Thai version (DRS-R-98) was used at the bedside to assess the presentation and severity of delirium on the evening of day 0 (pre-operative day), and twice a day for three days following surgery [29, 30]. The DRS-R-98-T has high sensitivity, specificity for delirium, and shows adequate interrater reliability [29, 30]. Prior to hospitalization, we recorded the use of anticholinergic medications, benzodiazepines, opiates, and psychotropic drugs, as well as relevant peri/post-operative clinical data such as operative time, blood loss, and the need for restraint due to psychomotor agitation. Post-surgery cardiovascular complications were registered including severe hypertensive state, atrial fibrillation, other arrhythmias, and acute coronary events. The body mass index (BMI) was computed as body weight (in kg) divided by height (in meter) squared.

The study protocol was reviewed and approved by the Faculty of Medicine, Chulalongkorn University, Bangkok, Thailand (registration number 528/61) institutional review board in accordance with the International Guideline for the Protection of Human Subjects, as required by the Declaration of Helsinki, The Belmont Report, CIOMS Guideline, and International Conference on Harmonization in Good Clinical Practice (ICH-GCP). All patients and their guardians (first degree family members) signed an informed consent form.

### Assays

Along with the clinical evaluation, venous blood samples were collected at 7 a.m., day 0. Blood samples were sent to the lab for IgA/IgG and cytokine/chemokine assays and frozen at -80°C until thawed. IgA- and IgG to diverse neuronal epitopes and HSP60 were assayed employing ELISA techniques MBP was purchased from Sigma-Aldrich® (St. Louis, MO, USA), and AQP4, MOG, NFP, GFAP, NMDAR, HSP60 were synthesized by Bio-Synthesis® (Lewisville, TX, USA). Alkaline phosphatase conjugated affinity pure goat anti-human IgA (α) and anti-human IgG, FCγ, were purchased from Jackson-Immuno Research® (West Grove, PA USA). Stock solution of 1 mg/mL was prepared from all antigens. From optimal dilution of 1:100-1:200, 100 µL of each antigen were prepared in 0.01M carbonate buffer pH 7.6, added to different wells of microtiter plates, and incubated overnight at 4°C. Plates were washed and 200 µL of 2% bovine serum albumin (BSA) in 0.01M PBS pH 7.4 was added to all wells in order to saturate the non-coated sites. After repeated washing, 100 µL of patients’ serum, calibrators and controls, at a dilution of 1:100 in 0.01 M PBS pH 7.4 containing 2% BSA and 0.05% Tween 20 were added to different duplicate wells and incubated for one hour at room temperature. Several wells containing all reagents, but not serum, were used for measurement of the reaction background or blank. During the incubation, sera antibodies bind strongly to antigen-coated microwells. Following washing and removal of unbound serum proteins, alkaline phosphatase labeled anti-human IgA at dilution 1:400, or anti-human IgG at dilution 1:800 was added to different sets of microwell plates, which were then incubated again for one hour. After a repeat of the washing step, addition of substrate para-nitrophenylphosphate concentration of 1 mg/ml, and incubation for 30 minutes at room temperature, a yellow color was developed in proportion to the concentration of antibody in the samples. Then 60 µL of 2N NaOH were added to stop the reaction and produce the endpoint color, which was measured by an ELISA reader at 405 nm. Antibody indices were measured using the following formula: antibody index = (OD of the sample – OD blank) / (OD of calibrator – OD of blank). Consequently, we computed two composite scores reflecting Gluta and NMDA receptor status and neuronal injuries as Ig responses to z GlutaR + z NMDAR (dubbed GlutaNMDAR) and z MBP + z MOG + z NFP + z AFAP (dubbed neuronal). As such, we entered 4 primary input variables in the analyses namely GlutaNMDAR, neuronal, AQP4 and HSP60.

As previously described [2], the Bio-Plex ProTM Human Chemokine Assays (Bio-Rad Laboratories, Inc. USA) were used to assay cytokines/chemokines, and samples were read with the Bio-Plex® 200 System (BioRad, Carlsbad, California, United States of America). The intra-assay CV for all analytes was 11.0%. We used fluorescence intensities, blank analyte subtracted, in the data analysis because fluorescence intensities are a better option than concentrations, especially when using multiple plates [2]. Because more than 20% of the measured concentrations of IL-2, IL-10, IL-12, and IL-13 were below the detection limit, these cytokines/growth factors were excluded from the analyses of single cytokines/growth factors. Nonetheless, these values were considered when creating immune profiles because detectable levels of those cytokines/growth factors may contribute to IRS/CIRS responses. The IRS and CIRS immune profiles, as well as the IRS/CIRS ratio, were the primary immune outcome variables in this study. The latter ratio was calculated as z (z IL-1 + z IL-6 + zTNF-α + z CXCL8 + z CCL3 + z sIL-RA + z IL-2 + z IL-12 + z IFN-γ + z IL-17) – z (z IL-4 + z IL-9 + zIL-13 + z IL-10) [2, 17].

### Statistics

Analysis of variance (ANOVA) was used to evaluate between-group differences in scale variables, whilst the X^2^-test was employed to determine connections between sets of categorical variables. The key outcome measures are the quantitative DRS-R-98 scale scores at days 2 and 3 being predicted by the biomarkers measured two days earlier. Moreover, we also extracted the first principal component (PC) of both DRS measurements (dubbed DRS2+3) to estimate the severity of delirium symptoms averaged over days 2 and 3. Moreover, we dichotomized the DRS2+3 scores using a cut-off value of 0.223 yielding a group with lower PC DRS2+3 and one with higher scores. The association of the IgA against self-antigens (input variables) and the dichotomized DRS2+3 scores (output variables) was examined using automatic logistic regression analysis. We computed the Odds ratio (OR) with 95% confidence intervals (CI) as well as parameter estimates (B with SE values) and used Nagelkerke values as pseudo-R^2^ effect sizes. We used generalized estimating equations (GEE) to examine the associations between DRS on days 2 and 3 (or the IRS/CIRS ratio at days 1 and 2) and the IgA- against self-antigens on day 0 while allowing for the intervening effects of baseline DRS, sex, age, previous neurological diseases, BMI, duration of surgery, time to surgery, estimated blood loss during surgery, and use of deliriogenic medications. Multiple associations between input and output variables were corrected using False Discovery Rate (FDR) p correction. The associations between the IgA reactivity to self-antigens and day 2 and 3 DRS scores, DRS day2+3 and the IRS/CIRS data on day 2 were computed using Pearson’s product moment correlation coefficients. Using the IgA values (and allowing for the effects of socio-demographic and clinical data) as input variables, automatic multivariate regression analysis was used to predict dependent variables (DRS-R-98 scores on days 2 and 3 and DRS2+3), while examining R^2^ changes, multivariate normality (Cook’s distance and leverage), multicollinearity (using tolerance and VIF), and homoscedasticity (using White and modified Breusch-Pagan tests for homoscedasticity). We utilized an automatic step-up procedure with a p-to-enter of 0.05 and a p-to-remove of 0.06. All these regression analyses were always bootstrapped using 5,000 samples, and these results are displayed if the findings did not agree. IBM SPSS Windows version 28 version 2022 was used for statistical analysis, and a significance level of 0.05 was used (two tailed tests). Given an effect size of 0.30, alpha of 0.05, power of 0.80, with five predictors, the a priori determined sample size for a multiple regression analysis is approximately 49 using G power analysis. The sample size for a repeated measurement design ANOVA should be around n = 60 when the effect size is 0.3, alpha is 0.05, and power is 0.80, with two groups and two repeated measurements (the biomarkers).

## Results

### Socio-demographic data

**Table 1** shows the socio-demographic, clinical and immune data in older adults divided into those with lower and higher peak DRS values. **Figure 1** shows the DRS values on day 0, day 1 and peak DRS scores in both patients with high and low peak DRS values. Repeated measurement ANOVA (age and CNS disease adjusted) showed a significant effect of time x group interaction (F=30.51, df=1/55, p<0.001), and time (F=6.78, df=1/55, p=0.012), and significant differences in DSR scores between both groups (F=17.75, df=1/55, p<0.001). People with higher peak DRS values were significantly older than the other subjects. There were no significant differences in sex, BMI, education, blood loss during surgery, time lag between fall and hospital admission and, length of stay in hospital until surgery between both groups. The total length of stay in the hospital was larger in patients with high peak DRS values. The IRS and IRS/CIRS composite scores were significantly higher in subjects with a higher DRS2+3 score. Electronic Supplementary File (ESF) Figure 1 shows the measurement of the DRS score from day 0 to day 3 (adjusted for age and T2DM).

**Figure 1.**
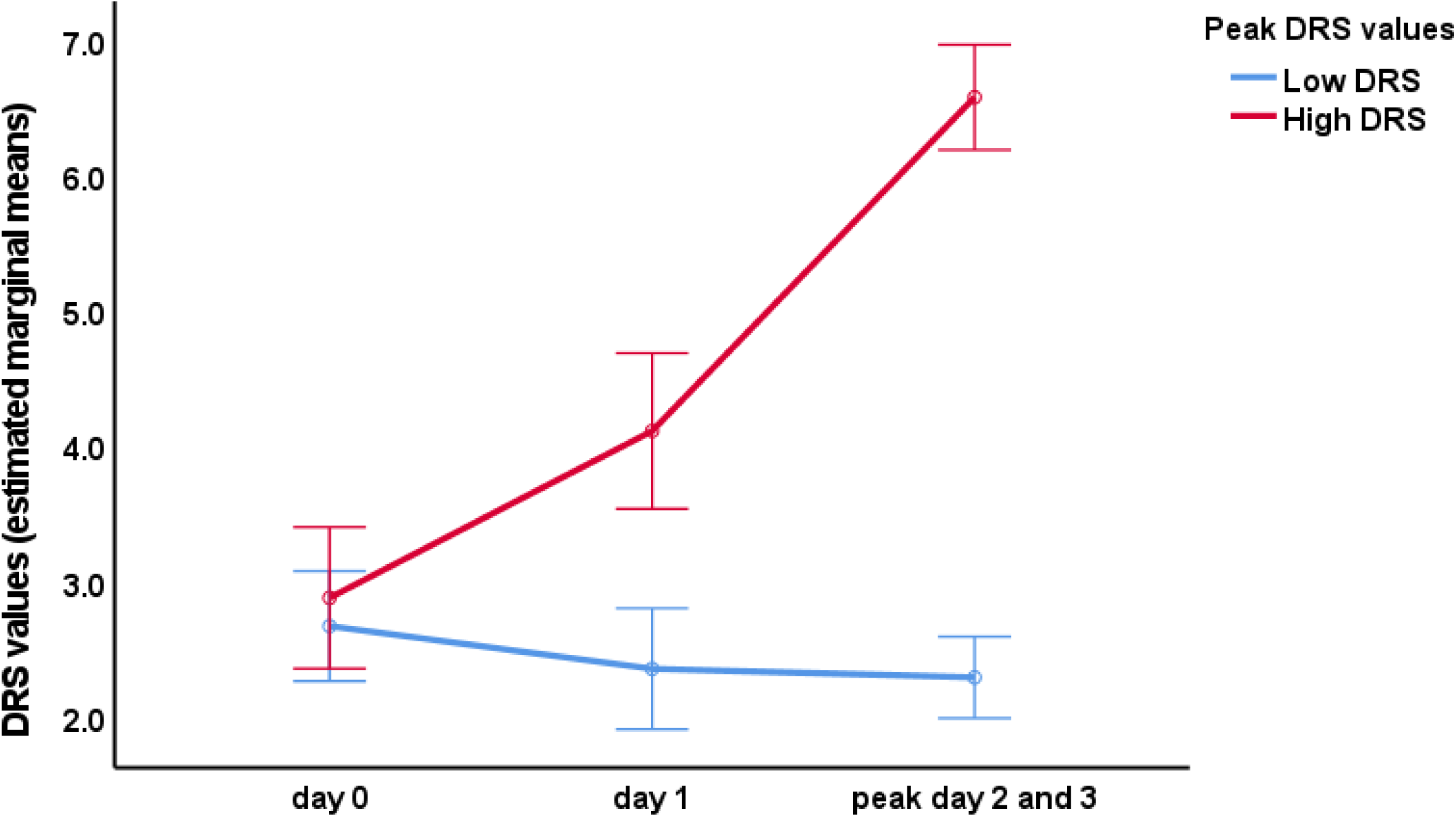
The Delirium Rating Scale, Revised-98-Thai version (DRS) scores on the surgery day (day 0) and 1, 2 and 3 days after surgery for hip fracture in patients with and without increased peak (day 2 and 3) DRS scores. In the latter, there is a significant effect of time (p<0.01), whilst no significant effect is established in the other patient group. The differences between both groups are significant on days 1 and 2 + 3 (p<0.01).

**Table 1.** Socio-demographic, clinical and immune data in older adults divided into those with lower and higher peak Delirium Rating Scale, Revised-98-Thai version (DRS) scores on days 2 and 3 (peak DRS2+3).

| Variables | Low peak DRS2+3<br>(N=36) | High peak DRS2+3<br>(N=23) | F/$\chi^2$ | df | P |
| --- | --- | --- | --- | --- | --- |
| DRS day0 | 2.28 (2.42) | 3.48 (2.50) | 3.36 | 1/57 | 0.072 |
| DRS day1 | 1.86 (1.94) | 4.87 (3. | 15.06 | 1/57 | <0.001 |
| Peak DRS day2+3 | 2.15 (0.95) | 6.78 (2.54) | 54.69 | 1/57 | <0.001 |
| Age (years) | 77.9 (7.7) | 85.0 (5.9) | 13.89 | 1/57 | <0.001 |
| Sex (F/M) | 28/8 | 18/5 | 0.00 | 1 | 0.965 |
| Education (years) | 8.3 (5.4) | 7.7 (6.4) | 0.18 | 1/57 | 0.676 |
| Body mass index (kg/m2) | 22.02 (3.13) | 21.51 (3.34) | 0.332 | 1/57 | 0.567 |
| Fall to hospital (hours) | 2.2 (4.9) | 1.3 (1.1) | 0.61 | 1/57 | 0.439 |
| Hospital to surgery (hours) | 74.4 (76.2) | 74.1 (58.5) | 0.00 | 1/57 | 0.989 |
| Length of stay in hospital (days) | 9.2 (4.4) | 12.1 (6.4) | 4.29 | 1/57 | 0.043 |
| Blood loss during surgery (mL) | 210.0 (115.6) | 201.3 (107.0) | 0.08 | 1/57 | 0.7734 |
| IRS (z scores) | -0.255 (0.938) | 0.424 (1.071) | 6.50 | 1/57 | 0.013 |
| CIRS (z scores) | 0.0423 (0.927) | 0.838 (1.064) | 0.03 | 1/57 | 0.876 |
| IRS/CIRS (z scores) | -0.258 (0.913) | 0.392 (0.966) | 6.69 | 1/57 | 0.012 |

Results are shown as mean (SD) or as ratios.: F: results of analyses of variance; $\chi^2$ : results of analysis of contingency tables
IRS: z unit-based composite score reflecting the immune-inflammatory responses system; CIRS: z unit-based composite score reflecting the compensatory immunoregulatory responses system

### Differences in IgA/IgG responses among DRS groups

**Table 2** shows the results of logistic regression analysis with the IgA antibody levels as input variable and the high DRS2+3 groups as dependent variable. We found that IgA neuronal, IgA GlutaNMDAR, IgA against AQP4 and HSP60 were significantly and positively associated with the high DRS2+3 group. These differences remained significant after FDR p-correction. Binary logistic regression analysis showed that IgA antibody levels to MPB (p=0.010), MOG (p=0.014), NFP (p=0.020), AFAP (p=0.016), GlutaR (p=0.023), and NMDAR (p=0.006) were significantly higher in the group with increased peak DRS scores. These differences remained significant after FDR p-correction. **Figure 1** shows the differences in the IgA values between both DRS groups. All differences were significant using ANOVA and the Kruskal-Wallis test. **Electronic file Figure 1** shows the mean IgG values to the same self-antigens, indicating that not one of those measurements was significantly associated with the DRS groups.

**Table 2.** Results of binary logistic regression (LR) and generalized estimating equation (GEE) analysis with the Delirium Rating Scale, Revised-98-Thai version (DRS), either as binary or scale variable, as dependent variables and Ig responses to self-antigens and immune data as explanatory variables.

| **Peak DRS2+3**<br>**Binary (LR)** | **B** | **SE** | **Wald (df=1)**<br>**(Nagelkerke)** | **p** | **OR** | **95% CI** |
| --- | --- | --- | --- | --- | --- | --- |
| IgA Neuronal | 0.915 | 0.370 | 6.10 (0.190) | 0.013 | 2.50 | 1.21; 5016 |
| IgA GlutaNMDAR | 0.885 | 0.345 | 6.57 (0.191) | 0.010 | 2.42 | 1.23; 4.77 |
| IgA Aquaporin | 0.692 | 0.304 | 5.19 (0.137) | 0.023 | 2.00 | 1.10; 3.63 |
| IgA HSP60 | 1.0612 | 0.403 | 6.94 (0.228) | 0.0081 | 2.89 | 1.31; 6.37 |
| **DRS day 2 and 3**<br>**(GEE, repeated**<br>**measurements)** | **B** | **SE** | **Lower 95% CI** | **Higher 95% CI** | **Wald (df=1)** | **p** |
| IgA Neuronal | 0.628 | 0.1666 | 0.302 | 0.955 | 14.23 | <0.001 |
| IgA MBP | 0.641 | 0.1673 | 0.313 | 0.969 | 14.70 | <0.001 |
| IgA MOG | 0.550 | 0.1804 | 0.196 | 0.903 | 9.29 | 0.002 |
| IgA NFP | 0.581 | 0.1573 | 0.273 | 0.890 | 13.67 | <0.001 |
| IgA GFAP | 0.649 | 0.1535 | 0.348 | 0.950 | 17.85 | <0.001 |
| IgA GlutaNMDAR | 0.616 | 0.1972 | 0.230 | 1.003 | 9.77 | 0.002 |
| IgA GlutaR | 0.542 | 0.1897 | 0.170 | 0.914 | 8.17 | 0.004 |
| IgA NMDAR | 0.637 | 0.1968 | 0.252 | 1.023 | 10.49 | 0.001 |
| IgA Aquaporin | 0.527 | 0.1967 | 0.142 | 0.912 | 7.18 | 0.007 |
| IgA HSP60 | 0.619 | 0.1498 | 0.326 | 0.913 | 17.10 | <0.001 |
| IRS | 0.425 | 0.1830 | 0.067 | 0.784 | 5.40 | 0.020 |
| CIRS | 0.201 | 0.2390 | -0.268 | 0.669 | 0.707 | 0.401 |
| IRS/CIRS | 0.426 | 0.1813 | 0.070 | 0.781 | 5.51 | 0.019 |
| **CV complications**<br>**Binary LR** | **B** | **SE** | **Wald (df=1)**<br>**(Nagelkerke)** | **p** | **OR** | **95% CI** |
| IgA composite | 0.117 | 0.049 | 5.79 | 0.016 | 1.13 | 1.02;1.24 |
| IgG composite | 0.111 | 0.056 | 3.97 | 0.046 | 1.12 | 1.01; 1.25 |

OR: Odds ratio; CI: confidence intervals
HSP60: heat shock protein 60; MBP: myelin basic protein; MOG: myelin oligodendrocyte glycoprotein; NFP: neurofilament protein; GFAP: glial fibrillary acidic protein; GlutaR: glutamate receptor; NMDAR: N-methyl-D-aspartate receptor
Neuronal: z composite score based on MBP, MOG, NFP and GFAP; GlutaNMDAR: z composite score combining GlutaR and NMDAR
IRS: z unit-based composite score reflecting the immune-inflammatory responses system; CIRS: z unit-based composite score reflecting the compensatory immunoregulatory system
CV complications: cardio-vascular complications following hip fracture surgery; IgA composite: z unit-based composite score comprising all IgA responses; IgG composite: z unit-based composite score comprising all IgG responses

### Associations between IgA/IgG values and the DRS scale scores

Table 2 shows the results of GEE analysis with the DRS scores on day 2 and the peak DRS scores as repeated measures and the IgA values as explanatory variables. The results show that the DRS scores from day 0 to 2-3 days later were significantly associated with the IgA levels to neuronal and GlutaNMDAR antigens HSP60, and AQP4. These differences remained significant after p correction. The solitary IgA levels to neuronal and GlutaNMDAR antigens were all significant and the strongest effects were found on HSP60 and GFAP.

**Table 3** shows the correlations between the 4 primary biomarker variables and the DRS scores on day 1, peak DRS2+3 scores and the actual changes in peak DRS2+3 scores after partialling out the effects of DRS day 0 (dubbed the residualized DRS2+3). There were no significant associations between the IgA values and DRS scores on day 1. There were significant associations between all IgA and these correlations remained significant after p correction.

**Table 3.** Intercorrelations between the Delirium Rating Scale, Revised-98-Thai version (DRS) scores, immune indices and IgA responses to self-antigens

| Variables | DRS day1 | Peak DRS2+3 | Res peak DRS2+3 | IRS day 1 | CIRS day 1 | IRS/CIRS day 1 |
| --- | --- | --- | --- | --- | --- | --- |
| IgA Neuronal | 0.034 (0.804) | 0.413 (0.002) | 0.469 (<0.001) | 0.374 (0.004) | 0.250 (0.063) | 0.327 (0.014) |
| IgA GlutaNMDAR | 0.036 (0.790) | 0.414 (0.002) | 0.457 (<0.001) | 0.417 (0.001) | 0.298 (0.025) | 0.340 (0.010) |
| IgA Aquaporin | -0.013 (0.924) | 0.330 (0.013) | 0.379 (0.002) | 0.406 (0.002) | 0.273 (0.042) | 0.342 (0.010) |
| IgA HSP60 | 0.056 (0.684) | 0.453 (<0.001) | 0.474 (<0.001) | 0.372 (0.005) | 0.260 (0.053) | 0.312 (0.019) |

DRS day 1: DRS score on day 1 post-surgery, peak DRS2+3: peak DRS scores on days 2 and 3 after surgery; res peak DRS: residualized changes in peak DRS2+3 scores after partialling out the effects of DRS day 0 (dubbed the residualized peak DRS2+3).
Neuronal: z composite score based on IgA responses to 4 neuronal self-antigens; GlutaNMDAR: z composite score combining IgA responses to the Glutamate and N-Methyl-D-Aspartate Receptors; HSP60: heat shock protein 60
IRS: z unit-based composite score reflecting the immune-inflammatory responses system; CIRS: z unit-based composite score reflecting the compensatory immunoregulatory system

There were no significant correlations between the IgA- and IgG values and age, sex, drug status, blood loss during surgery, smoking, and alcohol use (even without p correction). There were no significant correlations between the IgA- and IgG values to the same self-antigen (even without p correction). We found (see Table 2) that the cardio-vascular complications of surgery were in part predicted by the combined effects of IgA and IgG reactivity levels to self-antigens (entered a z unit-based composites based on the 8 self-antigens).

### Associations between immune data, DRS scores and IgA values

Table 2 shows the results of GEE analysis with the day 1 and peak DRS2+3 scores (repeated measurements) as dependent variables and the IgA antibody levels or IRS/CIRS data as predictors. Table 2 shows that the IRS and IRS/CIRS ratio, but not CIRS, significantly predicted the DRS scores from day 0 to days 2-3. Table 3 shows that the 4 key IgA values were all correlated with the IRS and IRS/CIRS, but not CIRS data.

### Results of multiple regression analysis

**Table 4** shows the results of multivariate regression analyses with the peak DRS2+3, DRS day 0, IRS/CIRS ratio, composite score IgA (computed as a z unit-based composite based on the 8 IgA responses to self-antigens), and length of hospital stay as dependent variables. Regression #1 shows that 40.6% of the variance in the peak DRS2+3 scores was explained by the regression on IgA to NMDAR and DRS day 0 indicating that the actual changes in DRS from baseline to 2-3 days later are associated with the IgA values. **Figure 2** shows the partial regression of the peak DRS2+3 score on IgA to NMDAR after adjusting for basal DRS values. We have rerun the same regression without the DRS day 0 values. Regression #2 shows that 36.0% of the variance in the peak DRS2+3 score was explained by the regression on IgA NMDAR age and CNS disease (all positively associated). Regression #3 shows that 25.9% of the variance in peak DRS2+3 scores was explained by age and use of selective serotonin reuptake inhibitors. The same table also shows the results of three regressions with the IRS/CIRS ratio as dependent variable. Regression #4 shows that 19.5% of the variance in the IRS/CIRS ratio on day 1 was explained by the regression on IgA MBP and CNS disease (all positively). We found (regression #5) that 29.1% of the variance in the IRS/CIRS ratio was explained by the combined effects of IgA to NMDAR, IgG to MBP and age (all positively associated). **Figure 3** shows the partial regression of the IRS/CIRS ratio on IgG MBP. Regression #6 shows that 23.1% of the variance in the IRS/CIRS ratio was explained by IgG- to MBP and age combined. There was a significant association between T2DM and increased composite IgA values with a low effect size (table 4). Length of stay in hospital was best predicted by the combined effects of CNS disease and peak DRS2+3 values.

**Figure 2.**
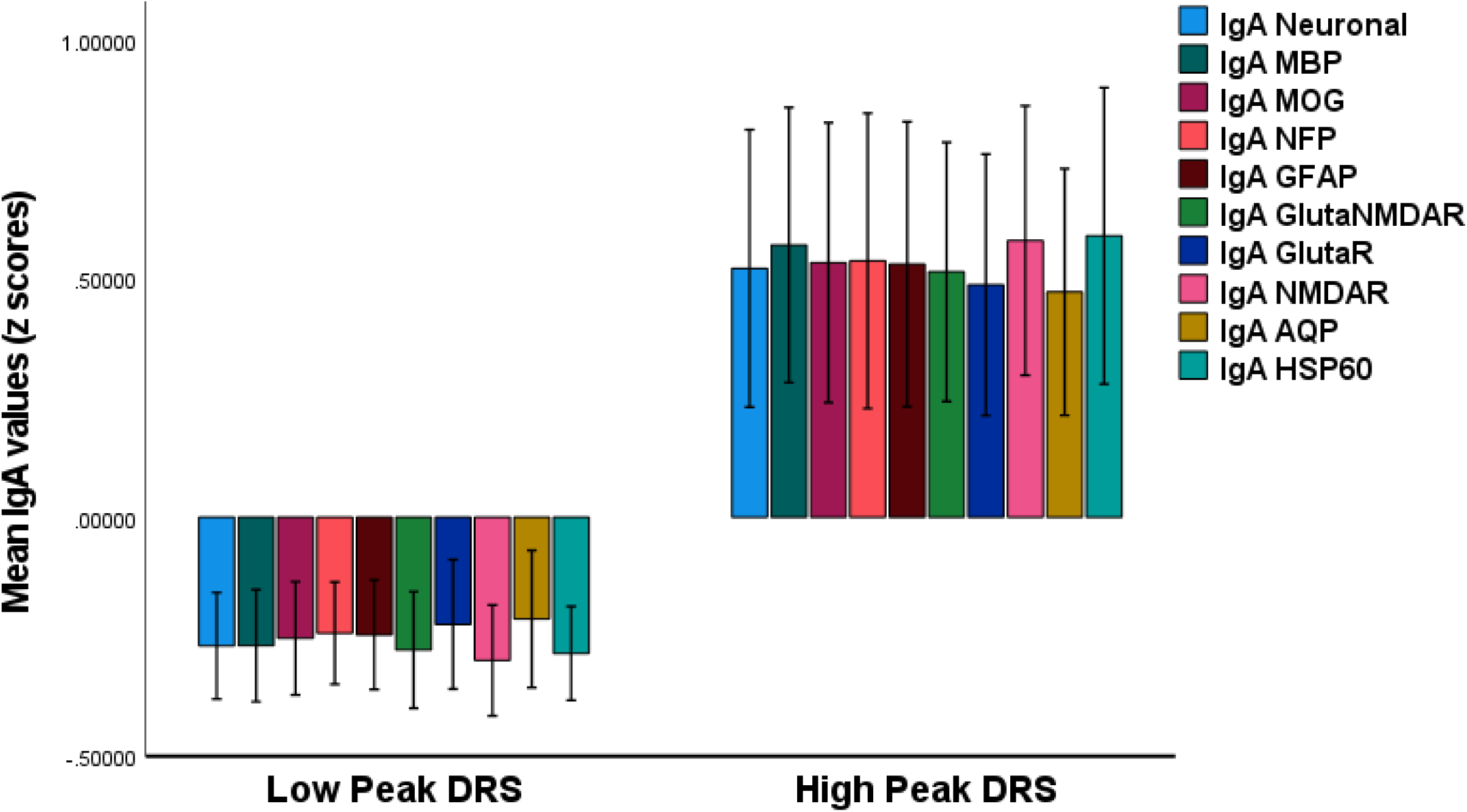
Clustered bar graph showing the IgA antibody levels to different neuronal tissue antigens and heat shock protein 60 (HSP60) in post-surgery patients with hip fracture with and without increased peak (day 2 and 3) Delirium Rating Scale, Revised-98-Thai version (DRS) scores. All IgA values are significantly different between both groups (p<0.01). MBP: myelin basic protein; MOG: myelin oligodendrocyte glycoprotein; NFP: neurofilament protein; GFAP: glial fibrillary acidic protein; GlutaR: glutamate receptor; NMDAR: N-methyl-D-aspartate receptor; IgA Neuronal: z composite score based on MBP, MOG, NFP and GFAP IgA values; IgA GlutaNMDAR: z composite score combining IgA responses to GlutaR and NMDAR

**Figure 3.**
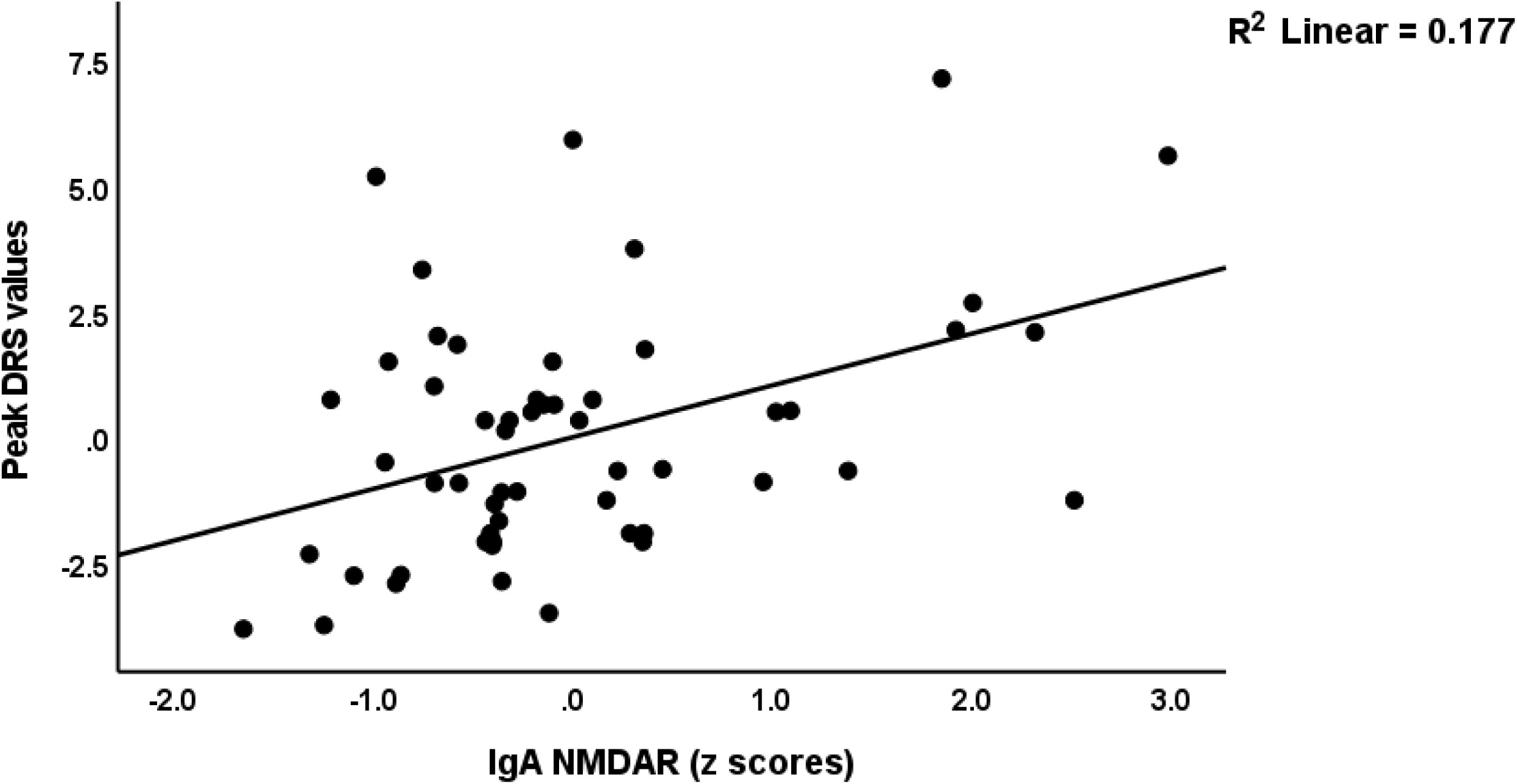
Partial regression of peak Delirium Rating Scale, Revised-98-Thai version (peak DRS) scores on the IgA antibody reactivity against the N-methyl-D-aspartate receptor (NMDAR).

**Figure 4.**
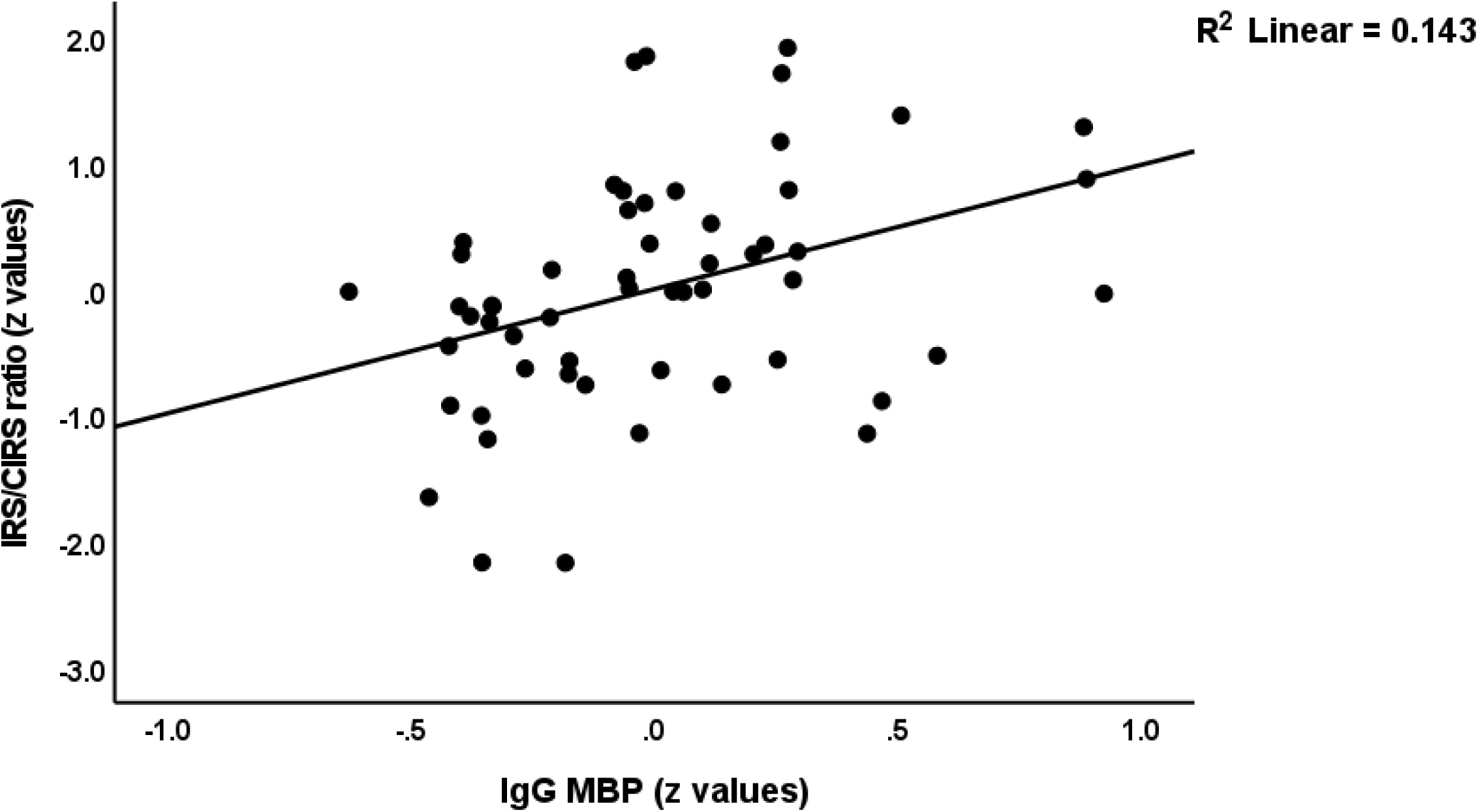
Partial regression of the IRS (immune-inflammatory response system) on CIRS (compensatory immunoregulatory system) ratio on IgG antibody reactivity to myelin basic protein (MBP).

**Table 4.**
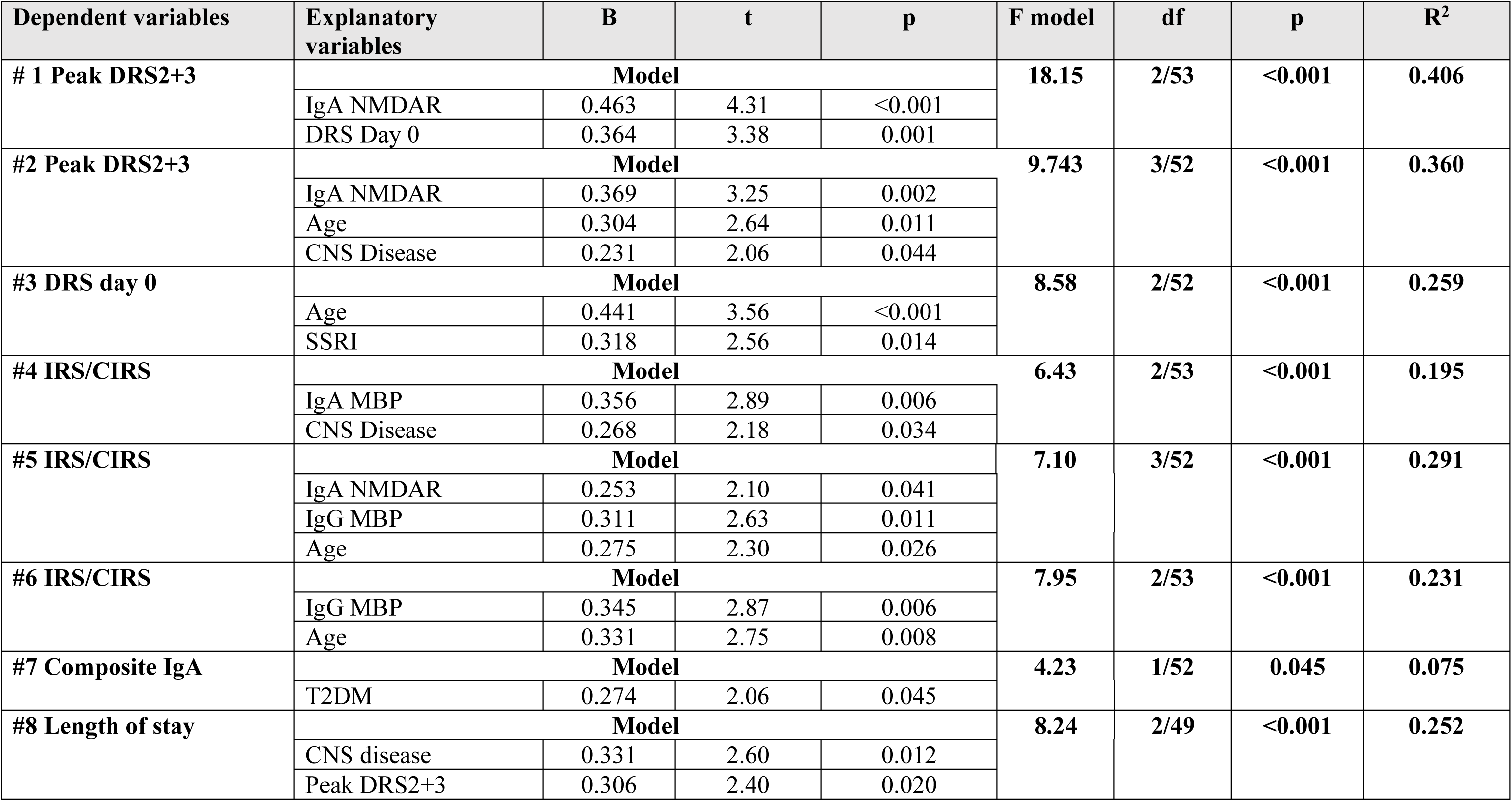
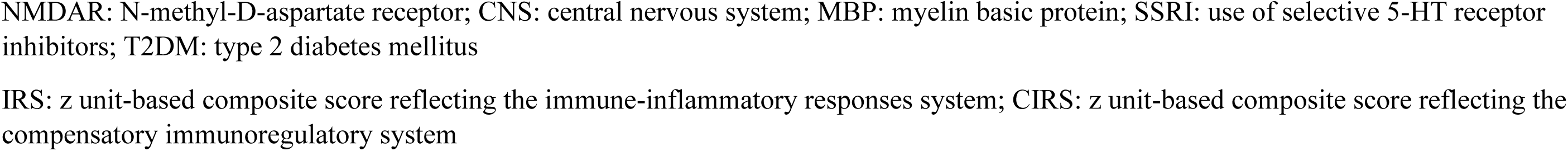
Results of multiple regression analyses with the peak Delirium Rating Scale, Revised-98-Thai version (DRS) scores on days 2 + 3 (peak DRS2+3) post-surgery or day 0, immune indices, a composite IgA response score and length of hospital stay as dependent variables

| Dependent variables | Explanatory variables | B | t | p | F model | df | p | R<sup>2</sup> |
| --- | --- | --- | --- | --- | --- | --- | --- | --- |
| # 1 Peak DRS2+3 | **Model** |  |  |  | **18.15** | **2/53** | **<0.001** | **0.406** |
|  | IgA NMDAR | 0.463 | 4.31 | <0.001 |  |  |  |  |
|  | DRS Day 0 | 0.364 | 3.38 | 0.001 |  |  |  |  |
| #2 Peak DRS2+3 | **Model** |  |  |  | **9.743** | **3/52** | **<0.001** | **0.360** |
|  | IgA NMDAR | 0.369 | 3.25 | 0.002 |  |  |  |  |
|  | Age | 0.304 | 2.64 | 0.011 |  |  |  |  |
|  | CNS Disease | 0.231 | 2.06 | 0.044 |  |  |  |  |
| #3 DRS day 0 | **Model** |  |  |  | **8.58** | **2/52** | **<0.001** | **0.259** |
|  | Age | 0.441 | 3.56 | <0.001 |  |  |  |  |
|  | SSRI | 0.318 | 2.56 | 0.014 |  |  |  |  |
| #4 IRS/CIRS | **Model** |  |  |  | **6.43** | **2/53** | **<0.001** | **0.195** |
|  | IgA MBP | 0.356 | 2.89 | 0.006 |  |  |  |  |
|  | CNS Disease | 0.268 | 2.18 | 0.034 |  |  |  |  |
| #5 IRS/CIRS | **Model** |  |  |  | **7.10** | **3/52** | **<0.001** | **0.291** |
|  | IgA NMDAR | 0.253 | 2.10 | 0.041 |  |  |  |  |
|  | IgG MBP | 0.311 | 2.63 | 0.011 |  |  |  |  |
|  | Age | 0.275 | 2.30 | 0.026 |  |  |  |  |
| #6 IRS/CIRS | **Model** |  |  |  | **7.95** | **2/53** | **<0.001** | **0.231** |
|  | IgG MBP | 0.345 | 2.87 | 0.006 |  |  |  |  |
|  | Age | 0.331 | 2.75 | 0.008 |  |  |  |  |
| #7 Composite IgA | **Model** |  |  |  | **4.23** | **1/52** | **0.045** | **0.075** |
|  | T2DM | 0.274 | 2.06 | 0.045 |  |  |  |  |
| #8 Length of stay | **Model** |  |  |  | **8.24** | **2/49** | **<0.001** | **0.252** |
|  | CNS disease | 0.331 | 2.60 | 0.012 |  |  |  |  |
|  | Peak DRS2+3 | 0.306 | 2.40 | 0.020 |  |  |  |  |

NMDAR: N-methyl-D-aspartate receptor; CNS: central nervous system; MBP: myelin basic protein; SSRI: use of selective 5-HT receptor inhibitors; T2DM: type 2 diabetes mellitus
IRS: z unit-based composite score reflecting the immune-inflammatory responses system; CIRS: z unit-based composite score reflecting the compensatory immunoregulatory system

## Discussion

### Increased IgA reactivity to neuronal antigens and HSP60 are associated with delirium

The first major finding of this study is that the IgA antibody reactivities to neuronal self-antigens, AQP4 and HSP60 strongly predict the severity of delirium symptoms a few days later. This is the first study to demonstrate such alterations in delirium; although, a few publications have indicated elevated serum/plasma concentrations of NFP, GFAP and MBP [22–25]. Previously, abnormalities in AQP4 expression were linked to cerebral edema and dehydration, both of which can produce delirium [27, 28]. Other results indicated that glutamate and the NMDAR system are linked to acute brain failure, including disinhibition and manic delirium [20, 21].

Our findings support the hypothesis that neuronal damage and abnormalities in the neuroaxis, AQP4, HSP90, and the glutamate/NMDAR system may contribute to the pathogenesis of delirium. Thus, elevated GFAP, MBP, and NFP levels are possible indicators of brain tissue and neuronal damage in brain trauma and neurological disorders [22–24]. For instance, people with traumatic brain injury, brain tumors, infections, stroke, and multiple sclerosis have elevated MBP levels [31,32,33]. MBP is an essential component of myelin sheaths and plays a crucial function in myelin formation; hence, it is referred to as the “executive protein of the myelin sheath” [34]. Following brain injury, MBP dissociates from the plasma membrane and is secreted into the extracellular matrix, where it functions as an antigen to activate immune-inflammatory pathways [34]. In addition, MBP may bind to the external surface of neuronal plasma membranes, causing neurotoxicity by disrupting lipid bilayers and increasing membrane permeability. As a result, MBP may have a role in demyelination following brain injury, as well as its repercussions, which include inflammatory reactions and cell death [34].

NFP is found in the cytoplasm of neurons, where it, in conjunction with microfilaments and microtubules, forms the cytoskeleton and modulates cell shape, axonal projections, and transport [35, 36]. NFPs are affected by neuronal trauma and neurodegeneration and may therefore be used as biomarkers of neuronal structural integrity and axonal fiber track damage [37]. Following brain trauma, NFP levels rise in CSF and plasma/serum and are detected in a variety of diseases, such as Alzheimer’s, Huntington’s, and Parkinson’s disease, multiple sclerosis, and toxic neuropathies [37]. In addition, the buildup of NFPs may trigger neurodegenerative processes, such as those observed in Alzheimer’s and Parkinson’s disease, amyotrophic lateral sclerosis, and diabetic neuropathy [38].

GFAP is an intermediate filament in the cytoskeleton of astrocytes and non-myelinating Schwann cells, and its expression is elevated during reactive astrogliosis in response to various CNS traumata [39, 40]. Following brain trauma, GFAP is released into the plasma via BBB breakdown, and elevated levels may therefore serve as biomarkers for acute CNS injuries, glial cell injury, and neurotoxicity [40]. Importantly, GFAP also serves as an autoantigen that provokes immune responses causing autoimmune responses in the CNS [40].

MOGs are an essential component of oligodendrocyte surface membranes and function as surface receptors, govern the integrity of oligodendrocyte microtubules, and play a crucial role in the formation and maintenance of myelin sheaths [41]. Upon translocation to the peripheral blood compartment, MOG may induce autoimmune reactions. Anti-MOG antibodies are found in multiple sclerosis and optic neuritis, however, it is unclear whether elevated antibody levels are part of an immune-inflammatory response or are pathogenic [42]. AQP4 may regulate osmotic water transport through epithelial cell and astrocyte membranes [43, 44]. AQP4-mediated water transport also promotes neuroexcitation and cell growth [45]. Antibodies to AQP4 may be generated as for example in atypical multiple sclerosis and MOG antibody-associated disease (MOGAD), which is an immune-inflammatory CNS disorder characterized by demyelination [42, 46].

The GlutaR and NMDAR are involved in the pathophysiology of acute brain failure for example during alcohol withdrawal, a lack of inhibition of NMDAR that may cause delirium [47], whist also increased glutamate activity is involved in the pathophysiology of delirium [48, 49]. Moreover, a significant association between delirium tremens and a functional polymorphism of the glutamatergic kainate receptor subunit GlurR7 was observed in patients with alcohol dependence [50]. Anti-glutamate receptor and anti-NMDAR antibodies are observed in serum in various conditions such as epilepsia, systemic lupus erythematosus, ataxia, encephalitis, stroke, trauma and neuropsychiatric disorders including schizophrenia [51–53]. Moreover, these antibodies may bind GlutaR and change glutamate-induced signaling and cause damage to brain neurons possibly leading to cell death and behavioral and cognitive impairments [51].

HSPs have protective activities against cell damage and are released into the blood under conditions of stress [54–58]. In several medical conditions, such as inflammatory illnesses, multiple sclerosis, trauma, cardiovascular disorders, diabetes mellitus, and sepsis, elevated levels of HSP have been found to damage the endothelium cells, leading to atherosclerosis [57,58,59]. Antibodies to HSP60 are linked to an increased risk of cardiovascular disease in older persons [56].

### The IgA- and IgG responses are associated with IRS activation

The second major finding of this study is that the IgG to MBP and IgA to neuronal self-antigens and HSP60 were substantially linked with IRS activation (a combination of M1, Th-1, and Th-17 profiles), but not with the CIRS composite index (computed as a composite of Th-2 and Treg functions). Anti-MBP IgG antibodies have hydrolytic, proteolytic, or antibody enzyme (abzyme) activity against MBP, thus leading to myelin sheath degradation and immune-inflammatory reactions [60, 61]. Moreover, IgA in general may also have pro-inflammatory effects, such as the breakdown of immune tolerance, the formation of complexed IgA, or the IgA-Fc alpha Receptor I axis [28,62,63,64]. As a result, these IgG and IgA responses may enhance the immune-inflammatory response and inflammation thereby increasing the risk of delirium in older adults [2]. Moreover, both the IgG- and IgA levels to neuronal tissue antigens and HSP60 predict cardio-vascular complications after surgery again suggesting that their pro-inflammatory properties may be involved.

### Interpretation of the results

The detection of IgA to MBP, GFAP, NFP, MOG, AQP4, and GlutaR in delirium may suggest that dysfunctions or injuries in the neuronal cytoskeleton, axons, astrocytes, oligodendrocytes, glial cells, and myelin sheath are associated with an increased risk for delirium. Notably, delirium was not associated with elevated IgG levels directed to the same self-epitopes. Nevertheless, natural PABs may be protective because they are a first-line defense against microbial invasion and have immunoregulatory, immune surveillance and homeostatic effects, and induce and maintain immune tolerance [28,64,65]. However, various conditions, such as toxic chemicals, mycotoxins, bacterial antigens, molds, and food antigens, can subvert their protective mechanisms, resulting in immune and B cell tolerance breakdown and PAB-related disorders [28, 64]. Importantly, PABs may elicit inflammatory responses thereby contributing to autoimmunity [28, 64]. In animal models, administration of such PABs to the brain may cause neuronal damage [66]. Such low-affinity natural PABs may act as precursors for the more pathogenic and high affinity Abs [67]. The data imply that elevations in IgA reactivity to a variety of neuronal tissue and HSP60 are risk factors for delirium [28], which along with the effects of the trauma and surgery, and the enhancement of the IRS contribute to the development of delirium symptoms in older adults.

### Limitations

The present study’s findings should be interpreted considering its limitations. First, the study was conducted on a smaller study group, and therefore, our results deserve replication. Second, the results would have been more interesting if we had also assayed the IgM responses to the same and other self-antigens, including oxidatively specific epitopes [68]. The latter have strong housekeeping and negative immunoregulatory effects [68] and their assays should shed further light on the mechanisms leading to delirium.

### Conclusions

IgA responses to MBP, MOG, NFP, GFAP, GlutaR, NMDAR, APQ4 and HSP60 strongly predict peak DRS scores at 2- and 3-days post-surgery. We discovered that IgA to NMDAR and baseline DRS scores explained 40.6% of the variance in peak DRS scores on days 2 and 3, and IgA to NMDAR, IgG to MBP, and age explained 29.1% of the IRS/CIRS ratio variance. Increased IgA to neuronal tissue antigens, AQP4, and HSP60 are risk factors for delirium. PAB-associated immune tolerance breakdown, as well as dysfunctions or injuries in the neuronal cytoskeleton, oligodendrocytes, glial cells, and myelin sheath, coupled with IRS enhancement and lowered CIRS protection can explain the onset of delirium at least partially.

## Data Availability

The dataset generated during and/or analyzed during the current study will be available from Prof. Dr. Michael Maes upon reasonable request and once the dataset has been fully exploited by the authors.

## Declarations

### Ethics approval and consent to participate

Approval for the study was obtained from the Institutional Review Board of the Faculty of Medicine, Chulalongkorn University, Bangkok, Thailand (registration number 528/61), in compliance with the International Guideline for Human Research protection, as required by the Declaration of Helsinki, was conducted according to Thai and international ethics and privacy laws.

Written informed consent was obtained before the study from the patients or their gardians (first-degree family members).

### Authors’ contributions

PT designed the conception, planned and collected the data, provided a delirium care, interpreted the results, drafted and revised the manuscript. YT planned and collected the data, provided a delirium care, revised the manuscript. ST planned and collected the data, provided an orthopedic care, revised the manuscript. SS prepared, analyzed and interpreted the laboratory specimens, revised the manuscript. MM designed the conception, analyzed and interpreted the data, drafted and revised the manuscript. All authors read and approved the final manuscript.

## Acknowledgements

We gratefully acknowledge the help of all psychiatry/orthopedic/anesthesiology nursing staff and residents involved in the execution of this study.

